# Covid-19 in children: is there any correlation with renal function and severity of the disease?

**DOI:** 10.1101/2020.10.20.20216440

**Authors:** Dedi Rachmadi, Ahmedz Widiasta, Hadyana Sukandar, Nanan Sekarwana, Dany Hilmanto

## Abstract

**Background:** Kidney manifestations are life-threatening conditions, such as end-stage renal disease (ESRD), especially when attributed to viral infections. The severe acute respiratory syndrome Coronavirus-2 (SARS-CoV-2), is an emerging health problem worldwide, potentially affecting all organs, including the kidney. Most reports on kidney manifestations were conducted mostly on the adult and elderly population, and limited on children. Therefore, this study aims to analyse the correlation between kidney manifestations with the renal function of pediatric patients suffering from COVID-19.

**Methods:** An observational analytic study was conducted in Hasan Sadikin General Hospital, Bandung, Indonesia, from March to August 2020. The demographic data, clinical signs, laboratory results, and notable kidney function were analysed, while the disease was classified as severe and nonsevere based on its clinical appearance. The Mann-Whitney test for nonparametric was used to analyze the collected data. *Results*. In this study, 16 COVID-19 children were selected as the research subjects, the median eGFR value in the severe group was lower (49.59 ml / minute / 1.73m2) compared to the nonsevere (113 ml / minute / 1.73m2), however, not statistically significant (p = 0.521). Significant high CRP and low thrombocyte levels were found in severe SARS-CoV-2 infection (p<0.05). *Conclusion*. A severe SARS-CoV-2 infection tends to affect the kidney, which is manifested as decreased glomerular filtration rate (GFR).

## Background

Since the emergence of severe acute respiratory syndrome Coronavirus-2 (SARS-CoV-2) in December 2019, it has haunted the world in the developed and developing countries due its fast spreading, therefore, in 2020 WHO declared the disease a pandemic. In Indonesia, COVID-19 was first reported on March 2, 2020, and treated at Hasan Sadikin Hospital, Bandung on March 17, 2020, which was a case of a 17-year-old girl that had close contact with her father that died of the disease. Furthermore, it is known to be caused by severe acute respiratory syndrome coronavirus 2 (SARS-CoV-2) (1), and leads to rapid activation of innate immune cells, especially in patients with severe infection. This novel disease primarily manifests as an acute respiratory illness accompanied by interstitial and alveolar pneumonia, however, it also affects multiple organs, such as heart, digestive tract, blood, central nervous system, and the kidney (2–5). The levels of various proinflammatory effector cytokines are elevated in COVID-19, especially in critically ill patients with acute kidney injury (AKI) (5,6).

Besides the respiratory system, the kidneys (urinary system) are the most often involved in the COVID-19 process (3,7–11). The various kidney involvement in children suffering from this disease is either mild, such as proteinuria or asymptomatic hematuria, to severe forms, such as acute kidney injury (AKI) (5,11,12).

This study aims to analyse the various characteristics of pediatric patients suffering from COVID-19, treated at Hasan Sadikin Hospital, Bandung, Indonesia, and to determine the involvement of their kidney function.

## Materials and methods

This was an observational and analytical study on pediatric patients suffering from COVID-19, treated at Hasan Sadikin Hospital in Bandung from March 13 to August 20, 2020. The research data was obtained from medical records after receiving clearance from the Health Research Ethical Committee, Hasan Sadikin Hospital, Bandung. The inclusion criteria for the research subjects were COVID-19 children with complete data on general characteristics, such as age, gender, treatment date, and underlying diseases. The clinical features and possible infection of the respiratory system were also noted. The laboratory and radiological features recorded include routine blood tests, serum urea, creatinine, CRP, and chest radiograph. The patients were classified into two groups based on the severity of the disease, namely, those hospitalized in the pediatric intensive care unit (PICU) as the severe, and unhospitalized as the nonsevere. The estimated glomerular filtration rate (eGFR) value was calculated based on the serum creatinine using the Schwartz formula. The data were presented descriptively using tabulations, which included the platelets, CRP, and eGFR, and were analyzed using different tests. When the data was normally distributed, the independent t-test was used, and when not normal, the Mann-Whitney method was utilized. The numerical data normality was analyzed using the Shapiro-Wilk test.

## Results

During the study period from March 13 to August 20, 2020, 16 children that showed positive PCR results for COVID-19 were recorded and included in this study. The resulting data was not normally distributed.

Demographic data and epidemiological characteristics of research subjects were presented in Table 1.

**Table 1.** Demographic and epidemiologic characteristics of children with COVID-19

| Characteristic | n |
| --- | --- |
| Age (years old) |  |
| < 2 years old | 7 |
| 2-6 years old | 2 |
| ≥ 6 years old | 7 |
| Median (range) : 5.16 (0.08 – 17) |  |
| Gender |  |
| Male | 6 |
| Female | 10 |
| Underlying disease |  |
| Healthy | 10 |
| Congenital | 5 |
| Tuberculosis | 1 |
| Epidemiologic history |  |
| Traveling history | 1 |
| Contact history with confirmed COVID-19 patient | 8 |
| Family cluster | - |
| Unknown | 7 |

Based on Table 1, the median age of the study subjects was 5.16 years, with a range between 27 days (0.08 years) and 17 years. More girls (10) than boys (6) was noted with a ratio of 1.67: 1. Out of the 16 study subjects, five had congenital abnormalities, i.e., two suffering from Tetralogy of Fallot, one with complete atrioventricular septal defect, and two others had intrahepatic cholestasis and jaundice accompanied by microcephaly and diaphragmatic hernia. Other epidemiological data showed that one patient was found to have travelled to the COVID-19 area, eight patients were known to have contact with an infected person, while seven had no contact at all.

### Clinical characteristics of children with COVID-19

The subjects in this study experienced various symptoms and signs as seen in Table 2.

**Tabel 2.** Clinical characteristics of children with COVID-19

| Characteristic | n |
| --- | --- |
| Clinical signs: |  |
| Asymptomatic | 6 |
| Fever | 4 |
| Cough | 3 |
| Breathless | 4 |
| Vomit/Diarrhea | - |
| Clinical classification: |  |
| Unclassified | 8 |
| URTI | 2 |
| Mild pneumonia | 3 |
| Critically ill cases | 3 |

Table 2 showed that, out of the 16 COVID-19 children, six did not express any symptoms, while the rest had fever, cough, or breath shortness. Meanwhile, based on the severity of the disease, three patients were critically ill and needed to be treated at the PICU, while others showed symptoms of upper respiratory tract infection or mild pneumonia.

### Laboratory X-ray and outcome of COVID-19 patients

From the 16 patients, the complete laboratory results included the hemoglobin, leukocyte, platelet, C-reactive protein (CRP), and serum creatinine values, which were converted to Estimated Glomerular Filtration Rate (eGFR) using the Schwartz formula. All the subjects were also examined for chest radiographs. Out of the 16 subjects, three were treated at the PICU room, and one died. The laboratory, radiological, and outcomes of the study subjects were shown in Table 3.

**Table 3.** Laboratoric and radiologic findings in children with COVID-19 according to disease severity

| Variable | Overall | Disease severity |  | p |
| --- | --- | --- | --- | --- |
|  |  | Nonsevere | Severe |  |
| I. Laboratoric findings: |  |  |  |  |
| Leucocyte (x10<sup>3</sup>/ mm<sup>3</sup>) | 10.64 (6.83-27.55) | 9.96 (6.83-27.55) | 10.64(10.40-22.87) | 0.573 |
| Lymphocyte (x10<sup>3</sup> /mm<sup>3</sup>) | 2.50 (0.42-11.57) | 2.72 (0.84-11.57) | 1.70 (0,42-2.74) | 0.161 |
| Thrombocyte (x10<sup>3</sup> /mm<sup>3</sup>) | 324 (87-589) | 370 (87-589) | 172 (124-173)<sup>*)</sup> | 0.049 |
| Hemoglobin (g/dL) | 11.4(9.8-24.5) | 12.2 (9.8-24.5) | 10.0 (9.8-11.8) | 0.161 |
| CRP (mg/L) | 0.46 (0.06-29.0) | 0.44 (0.06-0.97) | 19.94 (10.89-29.0)<sup>*)</sup> | 0.030 |
| eGFR (mg/dL) | 106.57 (27.93-315.74) | 113 (27.93-315.74) | 49.59 (35.60-243.97) | 0.521 |
| II. Radiologic findings : |  |  |  |  |
| Normal | 11 | 10 | 1 |  |
| Locally infiltrate | 3 | 2 | 1 |  |
| Bilateral infiltrate | 2 | 1 | 1 |  |
| Abnormal interstitial | - | - | - |  |
| III. Outcome : |  |  |  |  |
| Discharge | 2 | 2 | - |  |
| Pediatric ward | 11 | 11 | - |  |
| PICU | 2 | - | 2 |  |
| Death | 1 | - | 1 |  |

CRP, C-reactive protein, eGFR, estimated glomerular filtration rate, PICU, pediatric intensive care unit
Note: Data presented as median and range from minimum-maximum. The Mann-Whitney test was significant at $p < 0.05$

From Table 3, the platelet levels in the severe group were lower than those in the nonsevere (p = 0.049). There was no difference in hemoglobin and leukocyte values in the two groups (p> 0.05). Besides, the CRP results showed that these values were higher in the severe (19.94 mg/dl) compared to the non-severe group (0.04 mg/dl) with p = 0.030. Although the eGFR value in the severe group appeared to be lower (49.59 ml / minute / 1.73m2) compared to the nonsevere (113 ml / minute / 1.73m2), it was not statistically significant (p = 0.521). There was no data on the amount of urine or urinalysis results in the study subjects.

Radiological findings showed that 10 out of 13 patients that were not critically ill (nonsevere) and one of the three critical (severe) groups showed no abnormality. The radiological abnormalities of local or bilateral patches were seen in three out of 13 nonsevere and two out of three severe groups, while one of them died. There was no data regarding the results of the urinalysis of patients.

## Discussion

The subjects of this study were 16 children with their age ranging from 27 days to 17 years, seven patients aged less than two years old, and six under one years old. According to some studies, infants were rated more susceptible to COVID-19 infection (7,13). Therefore, they showed severe symptoms, especially with comorbidities, accompanied by decreased renal function, indicated by increased serum creatinine levels. In this study, the kidney function was absolutely different between the severe and nonsevere group. The subjects that experienced severe disease had a tendency of low eGFR, unfortunately, they were not statistically significant, probably due to the lack of sample size. Therefore, a power test was needed to achieve adequate sample size. The AKI occurred because in the clinically severe COVID-19 manifestation, several proinflammatory are being released, resulting in podocyte collapse, apoptosis, and even glomerular fibrotic (3,5,8,14).

Most of the patients experienced severe and critical illness or were hospitalized at the Pediatric Intensive Care Unit (PICU), and were diagnosed of comorbid diseases, such as hypertension, cardiovascular infection, or diabetes mellitus (3,8–11,15). In this study, the comorbid factors obtained were several congenital disorders, such as Tetralogy of Fallot, complete atrioventricular septal defect, and intrahepatic jaundice cholestasis with microcephaly, diaphragmatic hernia, and tuberculosis.

Data showed that COVID-19 was more lethal in men than women, with a mortality rate of 2.8% and 1.7%, respectively (16,17). Data from several European countries showed a similar number of cases between men and women, however, more severe in males (17). Mortality rate was higher in those with cardiovascular disease, and the reason this mechanism was rated more in women was still unclear (16,17). The mechanism and role of angiotensin converting enzyme2 (ACE2) and transmembrane serine protease2 (TMPRSS2) receptors, sex hormones’ role in responding to adaptive and natural (innate) immune responses, and a specific lifestyle regarding gender, health patterns, psychological stress, and socioeconomic conditions were investigated (1,16,17). The second protein for the invasion of SARS-CoV-2 into cells was TMPRSS2, which was mostly found in the prostate epithelium (1). The androgenic ligands and androgen receptors regulated the TMPRSS2 transcription (1). To enter cells, the SARS-CoV-2 virus binds to the ACE2 receptor and the TMPRSS2 cellular serine protease. ACE2 was a membrane-based protein expressed in various tissues, including the lungs and kidneys. Various reports have shown that the circulating ACE2 levels were higher in men than in women with kidney disease. The solute ACE2 was enzymatically active and had modest inhibitory effects on viral infection efficiency. However, there was no relationship between circulating ACE2 levels and COVID-19 involving epigenetic factors (5).

The regulation of angiotensin II receptor type I (AT1R) and renin activity by estrogen has been investigated. Estrogen modulated local RAAS in the atrial myocardium by downregulating ACE and simultaneously upregulating ACE2, AT2R genes encoding ACE2, and AT2R located on the chromosomes showing higher levels in women (17). However, various reports have shown that the pathological condition ACE2 was more common in men than in women (16).

A study showed that the hormone estrogen modulated the local renin-angiotensin-aldosterone (RAAS) system in atrial myocardial muscle by the down-regulation of ACE and simultaneously upregulating ACE2 and angiotensin II receptor type 2 (AT2R) (16,17). Interestingly, the genes encoding ACE2 and AT2R were located on the X chromosome, showing higher levels in females (16,17).

A prospective cohort study showed that women needed less ACEI doses to treat heart disease than men (16,17). The second protein for the invasion of SARS-CoV2 into cells was the surface serine protease TMPRSS2, found in cancerous prostate’s epithelium. However, they were also found in lesser amounts in the respiratory tract, although androgenic ligands and androgen receptors regulated the TMPRSS2 (1,7). The susceptibility and response to viral infection differed, in women, the circulating HIV viral RNA was 40% lower than that of men (16). Although, exposure to the influenza virus was higher in men, death was more common in women. The immune response to viruses varies with changes in sex hormone levels (((([17]. Most COVID-19 patients that were seriously ill or treated in Intensive Care had certain comorbid, such as hypertension, cardiovascular problems, or diabetes mellitus (7,9). In this study, five subjects had comorbid of congenital abnormalities, two with Tetralogy of Fallot, and one with Complete Atrioventricular Septal Defect, two had cholestasis jaundice intrahepatic with microcephaly and diaphragmatic hernia, and the other one with tuberculosis.

Several data that supported the role of gender in the incidence and severity of COVID-19 were mostly carried out in adults (16,17), meanwhile, it was limited in children. Three children experienced severe COVID-19 manifestation in this study, two of them were male, and this result matched theoretically. The mechanism that established the higher mortality rate for men than women was still unclear. Nonetheless, there was a suggestion for the role of the angiotension converting enzyme 2 (ACE2) receptor, the TMPRSS2 cellular serine protease, and sexual hormones in response to innate and adaptive immune responses. Tjeredore, to enter cells, the SARS-CoV-2 virus binds to the ACE2 and TMPRSS2 receptors (1). Various reports showed that the circulating ACE2 levels were higher in men than women with kidney disease (16–18). However, several preclinical trials have shown that the use of ACE2 had a protective effect against lung disease (19,20). However, presently, this role is still unclear when compared based on gender. A study conducted in China by Qiu et al. in March 2020, explained that, out of 36 children below 16 years that were infected with COVID-19, most of them were 3.8 years (21). In this study, the mean age in pediatric patients infected with COVID-19 was 5.16 years. The youngest age in this study was 27 days, and the oldest was 17 years. Another study conducted in Korea showed that out of 201 children with confirmed COVID-19, the youngest infected was 45 days, and the oldest was 15 years (3,18).

Some COVID-9 patients with AKI come to the hospital with complaints of vomiting and diarrhea. In these cases, there was an assumption that it was due to prerenal problem (8). In addition to these reasons, it was also suspected that the incidence was caused by an accompanying comorbid or direct invasion of the viral material into the kidney parenchyma (7,9). In this study, there were no vomiting and diarrhea complaints, which was one of the symptoms of AKI, although there were six patients with comorbid AKI not found. Interestingly, some postmortem histopathology findings indicated interference or damage to the kidney tissue of COVID-19 patients that were not previously detected through routine examination (increased levels of ureum or creatinine), indicating the possibility of subclinical AKI (11).

This study also found a significant higher C-reactive protein (CRP) and lower thrombocyte count in the severe group (Table 3). This condition probably correlated with the advanced cytokine storm process that involved in increasing proinflammatory mediators, such as interleukin-6 (IL-6), transforming growth factor-β (TGF-β), Tumor necroting factor-α (TNF-α), vascular endothel growth factor (VEGF), platelet derived growth factor (PDGF), IL-10, and soluble urokinase plasminogen activator receptor (suPAR) (5). Those proinflammatory and anti-inflammatory mediators released led to a disturbance in clotting cascades, while thrombus occurred in the later stages, and plasminogen stimulation with antithrombin-III activation took place in the fibrinolytic system (22). Therefore, fibrinolytic and fibrinogen substances were depleted, while clot formation and bleeding associated with disseminated intravascular coagulation (DIC) also occurred at the same time (23).

## Conclusion

Although children have less ACE2 receptor, the severe form of COVID-19 was found in their age. The severity of this disease was associated with kidney function, even in childhood age. Therefore, the pediatric nephrologist should be aware of kidney involvement for those with severe clinical manifestations.

## Data Availability

The data used to support the findings of this study are included in the manuscript.

http://www.dedirachmadi@yahoo.com

## Data Availability

The data used to support the findings of this study are included in the manuscript.

## Acknowledgments

This work was supported by the Academic Leadership Grant (ALG), Universitas Padjadjaran (contract no.2476/UN6.C/LT/2018)

